# Cochlear implantation restores cortical processing for spatial hearing in asymmetric hearing loss

**DOI:** 10.1101/2021.12.22.21268180

**Authors:** C. Karoui, K. Strelnikov, P. Payoux, A.-S. Salabert, C. James, O. Deguine, P. Barone, M. Marx

## Abstract

In asymmetric hearing loss (AHL), the normal pattern of contralateral hemispheric dominance for monaural stimulation is modified, with a shift towards the hemisphere ipsilateral to the better ear. The extent of this shift has been shown to relate to sound localisation deficits. In this study, we examined whether cochlear implantation to treat AHL can restore the normal functional pattern of auditory cortical activity and whether this relates to improved sound localisation. We recruited 10 subjects with a cochlear implant for AHL (AHL-CI) and 10 normally-hearing controls. The participants performed a voice/non-voice discrimination task with binaural and monaural presentation of the sounds, and the cortical activity was measured using positron emission tomography (PET) brain imaging with a H_2_^15^O tracer. The auditory cortical activity was found to be lower in the AHL-CI participants for all of the conditions. A cortical asymmetry index was calculated and showed that a normal contralateral dominance was restored in the AHL-CI patients for the non-implanted ear, but not for the ear with the cochlear implant. It was found that the contralateral dominance for the non-implanted ear strongly correlated with sound localisation performance (rho = 0.8, p < 0.05). We conclude that the restoration of binaural mechanisms in AHL-CI subjects reverses the abnormal lateralisation pattern induced by the deafness, and that this leads to improved spatial hearing. Our results suggest that cochlear implantation fosters the rehabilitation of binaural excitatory/inhibitory cortical interactions, which could enable the reconstruction of the auditory spatial selectivity needed for sound localisation.

## Introduction

In all species, one of the primary functions of hearing is to detect and locate brief sounds in order to direct visual attention and further analyse the sound sources (Heffner and Heffner, 1992). The ability to locate sound sources involves detecting stimulus disparities between the right and left ear in terms of the intensity (interaural level difference) and timing (interaural time difference) of the sounds. The information from the two ears is combined in a process known as binaural integration. This process enables spatial hearing as well as facilitating speech understanding in noisy environments. Studies have shown several advantages arising from binaural integration, including improved sound detection and identification in noise due to binaural redundancy, binaural unmasking and head shadow effects (Avan et al., 2015).

Asymmetric hearing loss (AHL), where there is profound deafness in one ear alone, is known to disrupt these binaural processes, leading to impairments in sound localisation and speech comprehension in noise. This has a significant negative impact on an individual’s quality of life (Nelson et al., 2019). The deficits can be seen when the hearing loss is bilateral, with one ear that hears better than the other, as well as when it is strictly unilateral. Studies have measured the extent of these binaural deficits, and in the case of sound localisation, this has been expressed as an increase in the angular error of localisation (Nelson et al., 2019) and as a right-left confusion in sound source lateralisation.

It is known that the information from each ear converges at an early level in the auditory pathway. When there is unilateral hearing loss, the binaural response properties of the auditory neurons are strongly affected, as revealed by electrophysiological recordings using animal models of congenital (Kral et al., 2013; Tillein et al., 2016) and acquired unilateral deafness (McAlpine et al., 1997; Popelár et al., 1994). In these studies, it has been found that the neuronal activity shifts towards the hemisphere ipsilateral to the normally-hearing ear, with both increased excitability/activity and a reduced latency. At the cortical level, the binaural responses display altered excitatory and inhibitory interactions (Kral et al., 2013; Tillein et al., 2016), which change the neuronal spatial properties (Barone et al., 1996; Clarey et al., 1992). These modifications induce profound functional reorganisation at the areal level, which affects the interhemispheric asymmetry of the auditory cortex (Ponton et al., 2001; Scheffler, 1998).

In patients with AHL, studies have shown that the auditory cortical asymmetry is altered, with a dominance shift from contralateral to ipsilateral with respect to the better-hearing ear (Ponton et al., 2001; Scheffler, 1998). This is therefore in line with the animal studies. In children with congenital AHL, this shift is accompanied by a weaker cortical representation of the deaf ear (Gordon et al., 2015). In a recent fMRI study on adults with acquired AHL (Vannson et al., 2020), the dominance shift was observed in the non-primary auditory cortex (NPAC), thus suggesting that the plasticity involves cortico-cortical interactions. Interestingly, the extent of this functional reorganization strongly correlated with the deficit in sound localisation. We hypothesise that this may be because the dominance shift reflects disruption to the sound field representation. This would be in line with the theory that the contralateral dominance seen in normally-hearing subjects relates to the representation of the contralateral sound field, rather than the contralateral ear input (Eisenman, 1974; Middlebrooks and Pettigrew, 1981; Phillips and Gates, 1982),

Studies on both humans and animals have clearly established that AHL has a deleterious effect on spatial hearing abilities, which is evident at both the behavioural and the neuronal levels (Gordon et al., 2015; Keating and King, 2013; Kumpik and King, 2019; Vannson et al., 2015). However, there is also evidence that some degree of auditory localisation is still possible, even when the binaural cues are altered. For example, following early unilateral deafness, sound localisation abilities can be developed using monaural spectral cue extraction (King et al., 2000; Slattery and Middlebrooks, 1994), specific training (Firszt et al., 2015), and following cochlear implantation (Arndt et al., 2011; Galvin et al., 2019; Távora-Vieira et al., 2015). These results clearly demonstrate the plasticity of auditory spatial processing and show that it can be achieved by means of adaptive strategies or by restoring binaural inputs to the brain (Keating and King, 2013).

The present study aims to explore the neural correlates of spatial hearing restoration following cochlear implantation for AHL, and more specifically, the lateralisation of brain activity in these patients. Ever since the seminal work of Van de Heyning et al. (2009), patients with AHL have been considered to be candidates for cochlear implantation (Arndt et al., 2011; Vermeire and Van de Heyning, 2009). Although the implants were initially used in these patients to target severe tinnitus, it was reasoned that the patients’ binaural abilities could also be improved (Arndt et al., 2011; Vermeire and Van de Heyning, 2009). This would involve combining the electrical cues conveyed by the implanted ear with the natural acoustic hearing in the other ear. It has since been found that the restoration of binaural integration is highly variable, presumably because of the difficulty involved in integrating the two types of information. Nevertheless, it has been found that the bilateral auditory input allows some degree of binaural hearing, as studies have shown improved sound localisation (Távora-Vieira et al., 2015) and speech recognition in noise (Arndt et al., 2020; Dorbeau et al., 2018; Galvin et al., 2019; Legris et al., 2018; Mertens et al., 2015). These improvements have been found to relate to various factors, including the pre-implantation performance, the hearing level of the non-implanted ear, the age at implantation, the duration of deafness and the age at the onset of deafness (Galvin et al., 2019; Mertens et al., 2015; Nelson et al., 2019; Távora-Vieira et al., 2015).

It is plausible that the restoration of binaural skills depends on an individual’s brain plasticity to enable the integration of both electrical and acoustical information. So far, with the exception of a few reports in children (Polonenko et al., 2018; Sharma and Glick, 2016), studies have not fully explored the functional brain responses in these patients. From the sparse existing literature, EEG studies have shown that the neural responses elicited by electrical and acoustical stimulation share similar latencies (Wedekind et al., 2020), but there is a weaker global auditory response (Legris et al., 2018) compared to normally-hearing controls, when both modalities are used.

The present investigation was motivated by the scarcity of studies on brain plasticity following cochlear implantation for AHL, and our interest in the recovery of spatial hearing. We used PET brain imaging to study the pattern of brain activation in response to natural auditory stimuli in cochlear implanted patients with AHL (AHL-CI); the cochlear implant precluded the use of other imaging techniques. We hypothesised that the stimulation of the deaf ear by a cochlear implant could restore, at least partially, the normal pattern of contralateral hemispheric dominance. Furthermore, as spatial hearing in AHL has been shown to relate to changes in hemispheric dominance (Vannson et al., 2020), we proposed that the restoration of contralateral dominance would relate to the restoration of spatial hearing.

## Materials and methods

### Subjects

Twenty subjects were recruited: 10 patients with a unilateral cochlear implant for AHL (AHL-CI) and 10 normally-hearing (NH) controls. The characteristics of the AHL-CI group are shown in Table 1. Patients were included if they had had a cochlear implant for at least 3 months in order to ensure a period of adaptation. For the non-implanted ear, the hearing thresholds ranged from normal (pure-tone average [PTA] <20 dB HL) to moderate hearing loss (PTA 50–70 dB HL); for the implanted ear, there was profound deafness (PTA >90 dB HL). All of the controls had normal thresholds over the range 0.5–4 kHz, with a PTA <20 dB HL, they were sex and age-matched to the AHL-CI group, thus giving a total of five women and five men in each group, with ages ranging from 46 to 74 years. Our study used a patient group and an imaging technique that are rare. The patients all had cochlear implants to treat asymmetric hearing loss; this is a group for whom rehabilitation evaluations are still exploratory; most patients treated so far have been included in research protocols. Our study also used H_2_^15^O PET imaging, an invasive and radioactive procedure, which imposes limitations on the number of subjects. It should be noted, however, that for PET H_2_^15^O activations studies, by accepting a Type 1 risk α of .05 with correction for multiple comparisons and a statistical power 1 – β of .90, the sufficient sample size per group is estimated as 10-12 subjects.(Andreasen et al., 1996)

**Table 1.** Characteristics of the subjects with a cochlear implant (CI)

| Code | Age<br>range<br>(years) | Sex | Aetiology | Duration<br>of HL<br>(years) | CI<br>experience<br>(years) | CI<br>Manufacturer | CI<br>side | Contralateral<br>pure tone<br>average |
| --- | --- | --- | --- | --- | --- | --- | --- | --- |
| **P5** | 61-70 | M | PHLEBITIS | 2 | 3 | Advanced<br>Bionics | R | 37 |
| **P7** | 51-60 | F | TRAUMA | 10 | 4 | Cochlear | L | 16 |
| **P8** | 51-60 | M | SUDDEN | 33 | 3 | Oticon<br>Medical | R | 27 |
| **P10** | 61-70 | F | SUDDEN | 9 | 3 | Cochlear | R | 15 |
| **P11** | 51-60 | F | OTOSCLEROSIS | 22 | 2 | Med-El | L | 36 |
| **P12** | 41-50 | M | OTOSCLEROSIS | 13 | 4 | Cochlear | L | 57 |
| **P13** | 71-80 | F | SUDDEN | 8 | 4 | Cochlear | L | 65 |
| **P14** | 71-80 | F | SUDDEN | 1 | 0.8 | Advanced<br>Bionics | R | 32.5 |
| **P16** | 51-60 | M | Menière's disease | 20 | 0.4 | Cochlear | L | 68 |
| **P17** | 51-60 | M | Menière's disease | 5 | 0.3 | Med-El | L | 50 |

The study was approved by the French South West and Overseas ethics committee (approval number: 2016-A01442-49) and conducted according to the Declaration of Helsinki including the signed by all participants informed consent form before taking part in the study.

### Localisation task

Among the different audiological evaluations, we selected horizontal localisation performance, which reflects more directly the level of binaural integration. An array of 12 loudspeakers was set up in an anechoic chamber, each at a distance of one metre from the subjects, with an angular separation of 15° (Chan et al., 2008) and located behind the subjects’ ears at the same height. The stimulus was a broadband impulse presented at 65 dB SPL. The subjects were asked to indicate where the noise had come from by clicking on one of the loudspeakers on the computer screen. For each run, there were 24 trials where the stimulus was presented randomly from one of the loudspeakers, with two presentations per loudspeaker. To minimize the use of loudness cues for the localisation task, the amplitude of the stimulus could randomly increase or decrease by 6 dB from trial to trial (roving) (Galvin et al., 2019). The AHL-CI group completed the localisation test under four different conditions: with the cochlear implant activated (CI-ON) or deactivated (CI-OFF), with a roving amplitude of 6 dB from trial to trial, and without a roving amplitude (non-roving). The NH group was only assessed using the roving condition. The localisation accuracy was reported as the average root mean square (RMS) error in degrees.

### PET procedure

The subjects were scanned in a shielded, darkened room. The head was immobilized and aligned transaxially to the orbitomeatal line using a laser beam. Regional distribution of radioactivity was obtained using a Biograph 6 TruePoint HiRez (Siemens Medical) PET scanner with full volume acquisition. The duration of each scan was 80s; approximately 6 mCi of H_2_^15^O was administered to each subject. A measure of auto-attenuation correction was obtained by performing a CT scan before the PET H_2_^15^O.

### Sound stimuli during the PET scan

All of the stimuli were taken from our database developed in previous studies (Massida et al., 2011; Vannson et al., 2020). Subsets of 500ms-long vocal and non-vocal stimuli were randomly presented using PsychoPy2 (Peirce et al., 2019). Vocal stimuli were 55 different sounds, including speech sounds (words in non-French languages and non-semantic syllables) and non-speech sounds (e.g., laughs, coughs). Non-vocal stimuli were different environmental sounds (alarms, car horns, bells …). The subjects were given a two-alternative forced choice task to indicate whether they heard a vocal or a non-vocal sound by clicking on the left or right button of a mouse. The duration of each trial was determined by the subject’s response time, but this was limited to a maximum of 8 seconds.

During the PET scan, the subjects had their eyes covered, and the auditory stimulation began at the descending part of the bolus curve, 10–20 seconds before the tracer reached the brain. For the NH group, the stimuli were delivered using inserts; for the AHL-CI group, the sound was delivered either directly via a cable connected to the implant, or via an insert in the non-implanted ear (after amplitude correction for hearing loss), or via the two systems simultaneously. There were four different conditions: (1) a baseline condition, where the subjects were instructed to lie quietly in the scanner, (2) a binaural condition, where the sounds were presented to both ears, (3) a right-ear monaural and (4) a left-ear monaural conditions. There were two runs for each condition, each lasting 80 seconds, the order of the runs was randomised for each subject, as was the order of the trials within each run. The total duration of the PET scan was 80 minutes.

### PET analysis

The PET data were analysed using SPM12 software and included realignment and spatial normalisation to the Montreal Neurological Institute template. The images of the patients with a right-sided CI (four patients) were flipped so that all of the implants were on the left side in the final images. The global signal was normalised within and between the subjects. The analyses focused at the early cortical levels (primary [PAC] and non-primary [NPAC] auditory cortex) based on our findings on UHL patients (Vannson et al., 2020); whole-brain analyses will be presented in a separate article. We used the combined cytoarchitectonically defined auditory areas Te1.0, Te1.1, Te1.2 and Te3.0 as the regions of interest, developed and implemented in the SPM Anatomy Toolbox (Eickhoff et al., 2006). We used the linear mixed-effects model (“lme4” package in R) with repeated measures for the comparisons between groups and conditions.

An asymmetry index (AI) of the brain activity in the auditory regions was calculated to study the lateralisation pattern of auditory activity with respect to the baseline: AI = (Contralateral - Ipsilateral) / (Contralateral + Ipsilateral). Contralateral and ipsilateral refer to the cortical side contralateral and ipsilateral to the stimulated ear.

In addition, the respective influence of the contralateral and ipsilateral ear on the auditory activity in each cortical hemisphere was estimated using the aural preference index (API): API = (Contralateral ear – Ipsilateral ear) / (Contralateral ear + Ipsilateral ear), where contralateral refers to the activity elicited by the contralateral ear and ipsilateral refers to the activity elicited by the ipsilateral ear. For each auditory cortex, the values are positive in the case of contralateral aural preference and negative in the case of ipsilateral aural preference.

## Data availability

The data that support the findings of this study are available from the corresponding author, upon reasonable request.

## Results

### Auditory test performance

The participants were given a voice/non-voice discrimination task during the PET scan. Performance was assessed by calculating the hit rate under each condition. Using a linear mixed-effects model with the group and the condition as fixed effects, significant results were found for the group, the condition and the interaction. Overall, the AHL-CI patients had poorer scores than the controls, with an average hit rate of 63±27%(SD), compared to 93±6% for the controls (p < 0.01; Figure 1A). Although the patients’ performance depended on the condition (binaural stimulation, acoustic stimulation or cochlear implant [CI] stimulation), it remained above chance (bootstrap, p < 0.05). The AHL-CI patients’ hit rates for the CI-only condition (49±27%) were significantly lower than for the binaural and acoustic conditions (77±19% vs 68±28% respectively, p < 0.01). We observed a weak difference between the binaural and acoustic conditions (p < 0.05), suggesting that the information provided by the implant slightly improved global auditory performance. The level of residual hearing, (PTA values), correlated with the scores for the binaural (r = 0.61, p < 0.01) and the acoustic (r = 0.63, p < 0.01) conditions.

**Figure 1.**
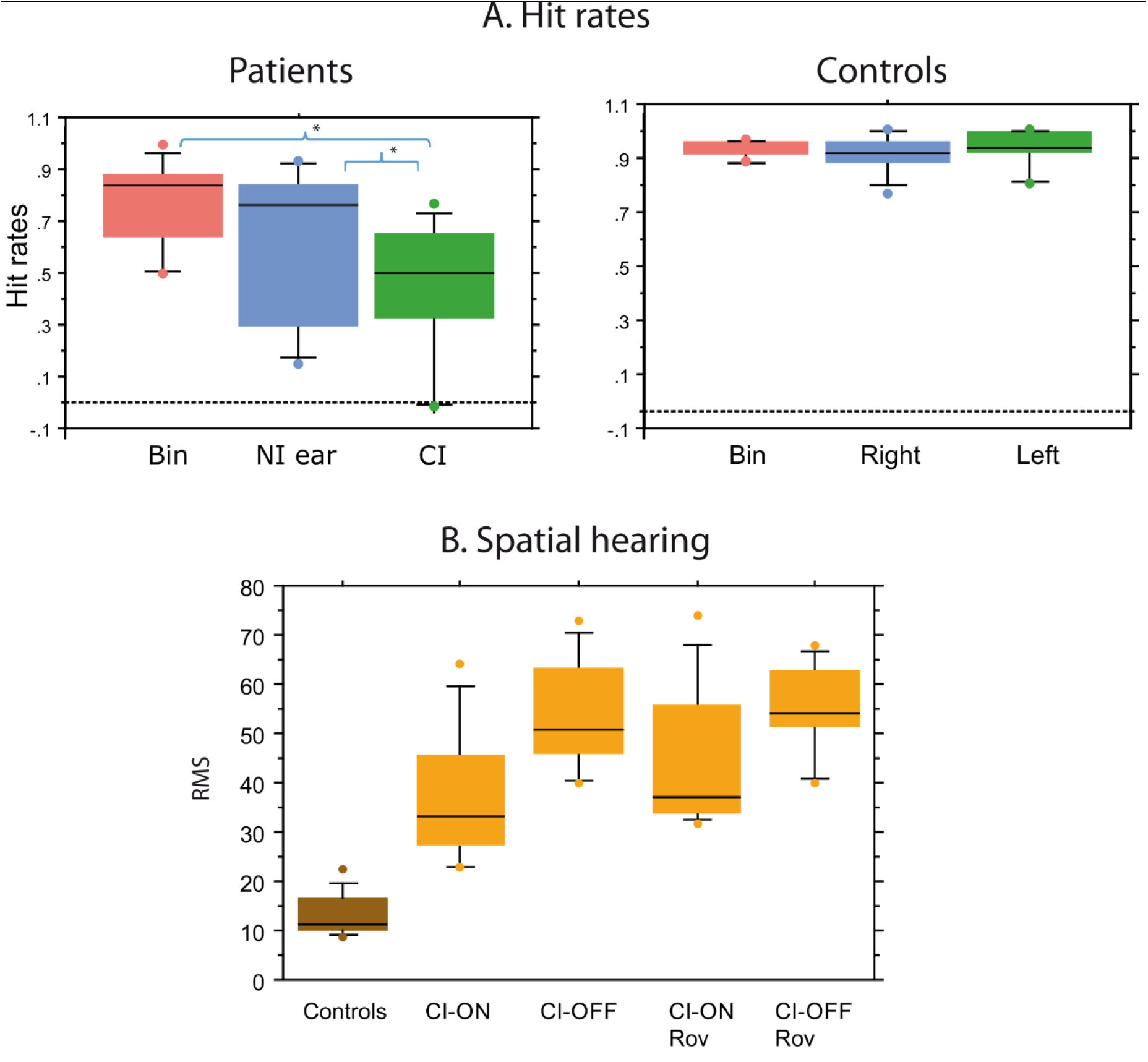
Hit rates on the voice/non-voice discrimination task and spatial hearing scores. Bin – binaural condition. NI ear – non-implanted ear. CI – cochlear implant stimulation. RMS – average root mean square error in degrees. A. Patients had significantly lower hit rates than the controls for all of the stimulation conditions; their hit rates for the CI condition were significantly lower than for the binaural and acoustic conditions. A small difference was observed between the binaural and acoustic conditions in the patients. B. Patients made significantly more errors during the spatial perception task than the controls. Without amplitude roving, there was a significant difference between the CI-ON and CI-OFF conditions, showing that the implant improved spatial localisation. However, when amplitude roving was introduced, there was only a trend towards better performance in the CI-ON condition (p = 0.08).

The spatial hearing abilities were evaluated just before the PET scan using a free-field sound localisation task. Performance was assessed with the cochlear implant on (CI-ON; i.e., a binaural, bimodal condition) and with the cochlear implant off (CI-OFF; i.e., an acoustic, monaural condition). On average, the AHL-CI subjects had poorer sound localisation than the controls (p < 0.05; Figure 1B). This was observed, as expected, in the CI-OFF condition, where the RMS errors often exceeded the range of 50–60°. When the amplitude of the sounds was constant, performance was significantly better in the CI-ON condition than in the CI-OFF condition (paired t-test; p = 0.01); in the more challenging roving amplitude condition, there was only a trend towards better performance in the CI-ON condition (p = 0.08). However, for this latter condition, seven of the ten subjects had superior sound localisation in the CI-ON condition, with a binaural benefit ranging from +9 to +33%. Thus, there was a beneficial effect, albeit limited, of the implant on spatial hearing, similar to the results found for discrimination task.

### PET results

#### A. Average activity in the auditory areas

The auditory activity in the PAC/NPAC regions was assessed for the different stimulation conditions (Figure 2A & B). A linear mixed effects model was used to analyse the differences between the subject groups, the stimulation conditions and the side of the auditory cortical activity (contralateral and ipsilateral to stimulation). The results showed significant differences for all three (Table 2, Figure 2A), the activity in the auditory regions being reduced by about 5% in the AHL-CI subjects compared with the controls (group effect; p < 0.01). During acoustic stimulation, there were higher levels of activity in the hemisphere contralateral to the stimulation, both in the AHL-CI patients and the controls (side effect; p < 0.05). In contrast, during CI stimulation, the activity was higher in the hemisphere ipsilateral to the implanted ear (condition: side interaction, p < 0.01), and was found to be higher than the ipsilateral activity during acoustic stimulation (post-hoc comparisons p < 0.01). The opposite pattern was found for contralateral activity: the auditory activation levels were higher for the acoustic than for the CI stimulation (p < 0.01). Altogether, these results suggest that the implant only partially restores a normal pattern of contralateral auditory cortical activation.

**Table 2.** The effects of subject group, stimulation condition and hemispheric side on auditory cortical activity.

| Effects and interactions | p |
| --- | --- |
| Condition | 0.00016 |
| Side | < 2.2e-16 |
| Group | 0.024 |
| Condition:Side | 0.002 |
| Condition:Group | 0.016 |
| Side:Group | 0.0003 |

**Figure 2.**
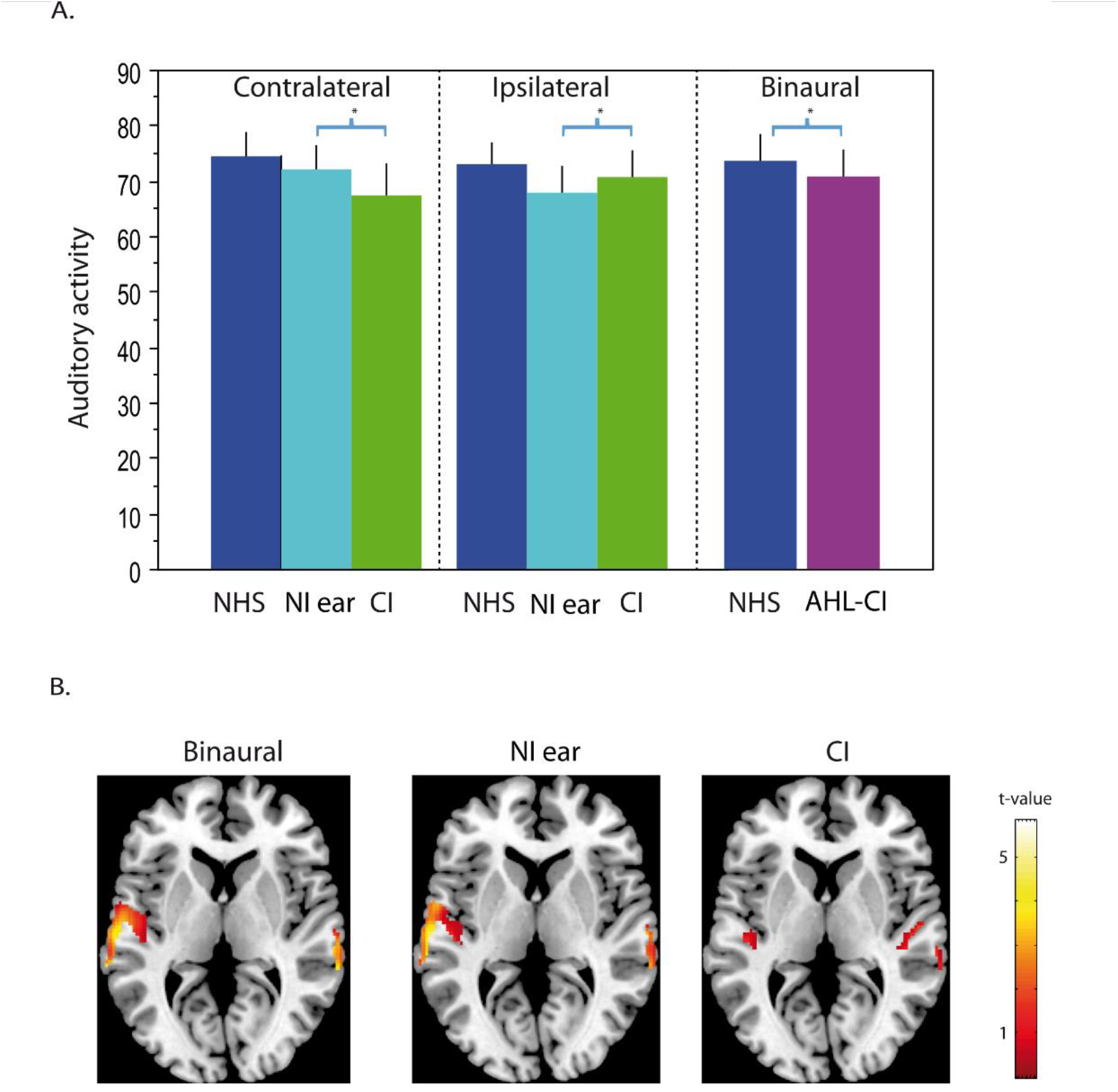
Auditory cortical activity in patients and controls. **A**. Contralateral and ipsilateral sides are indicated with respect to the stimulated ear. AHL-CI –asymmetric hearing loss with cochlear implants. NHS – normally-hearing subjects. NI ear – non-implanted ear (patients). CI – cochlear implant stimulation (patients). For the binaural bar chart, the average of both auditory cortices is presented. AHL-CI subjects had significantly lower activity in the auditory regions than the controls. In both patients and controls, during acoustic stimulation, activity was higher on the side contralateral to the stimulation. However, during CI stimulation in patients, activity was higher in the ipsilateral than the contralateral auditory cortex. **B**. Illustration of activity in patients during binaural stimulation, better acoustic ear stimulation (right ear for this image) and CI (left side) stimulation. It can be seen that the activity during binaural stimulation is similar to the acoustic stimulation.

We carried out further analyses to determine whether the auditory cortical activity relates to the level of residual hearing in the non-implanted ear. Significant negative correlations with PTA (p < 0.01; Figure 3) were observed, but only for the auditory cortex contralateral to the acoustic stimulation. This was found for both the binaural and the acoustic conditions (r = 0.62 and 0.52, respectively; Figure 3A and B), and it implies that the level of hearing loss negatively impacts the level of auditory cortical activity. We also found a similar negative correlation when the cochlear implant was stimulated, but this was for the cortex ipsilateral to the implant (r = -0.54, p < 0.01; Figure 3C). This suggests that the level of hearing loss affects the overall cortical excitability and influences the response to peripheral stimulation, whether acoustic or electrical.

**Figure 3.**
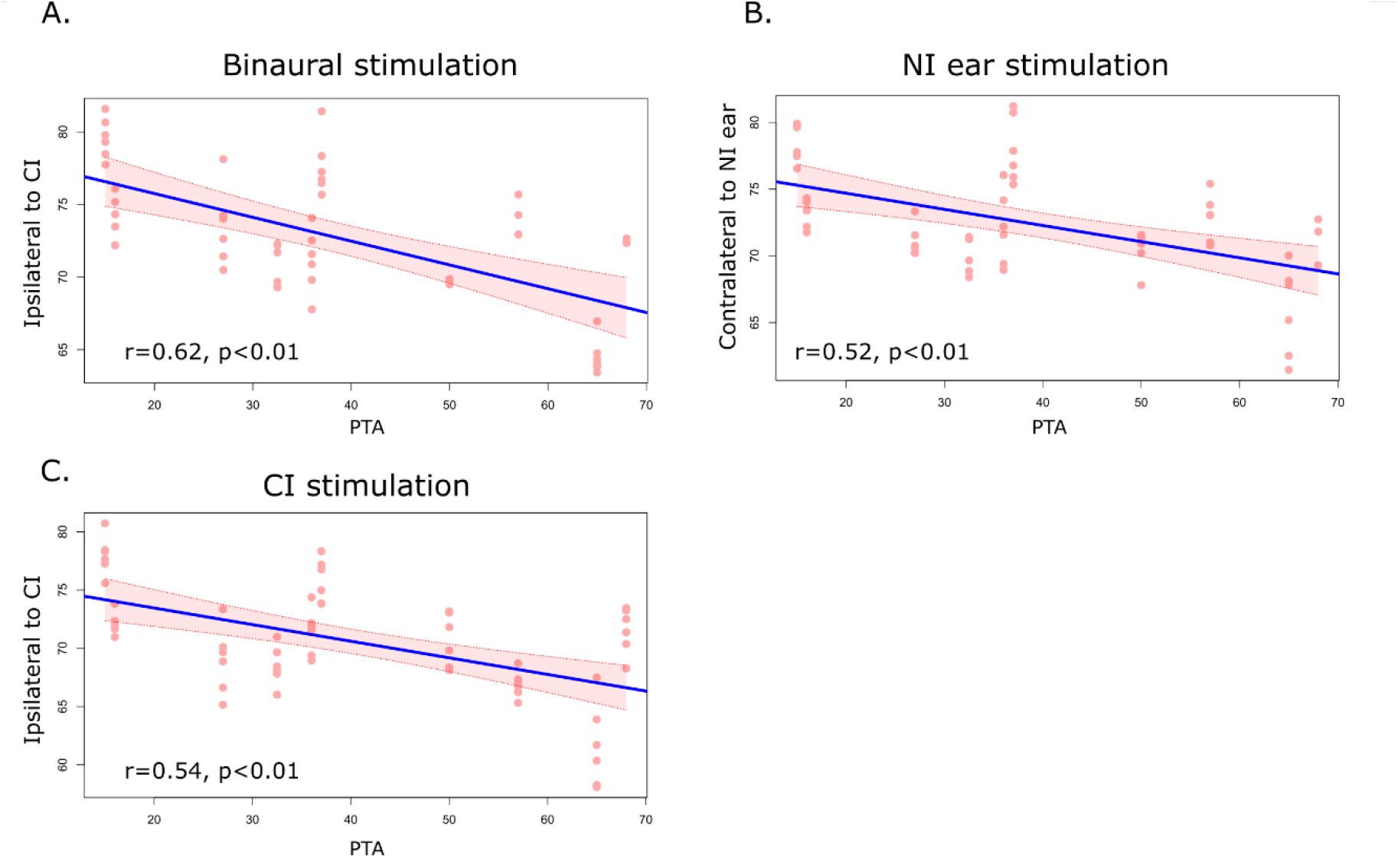
Auditory cortical activity and deafness. NI ear – non-implanted ear. CI –cochlear implant. PTA – pure tone average. Significant negative correlations were only observed between the PTA and activity in the auditory cortex contralateral to the acoustic ear (i.e., ipsilateral to CI).

We also carried out analyses to determine whether the duration of the hearing loss relates to the cortical activity. We found a weak, negative correlation for the activity contralateral to the implant during binaural stimulation (r = -0.36, p < 0.01) and CI stimulation (r = -0.37, p < 0.01), but not during the acoustic stimulation (r = -0.13, p = 0.93). This implies that a longer duration of deafness weakens the restoration of auditory activity in the hemisphere contralateral to the implant.

The influence of post-implantation experience was also assessed. The duration of CI experience correlated with activity in the cortex contralateral to the implant during binaural (r = 0.32, p < 0.05) and acoustic stimulation (r = 0.36, p < 0.01), but not during the CI-only condition (p = - 0.08, p = 0.55). No significant correlations were found for the ipsilateral cortex.

To summarise, the auditory cortical activity was found to be lower, on average, in the AHL-CI subjects than in the controls. This can be attributed to weaker activation from the cochlear implant as well as hearing loss in the non-implanted ear.

#### B. Auditory activity and test performance

For the AHL-CI patients, we examined whether the auditory activity was related to performance on the sound discrimination task (hit rates). Correlation analyses were run and revealed several significant positive correlations. These were found for the auditory cortex contralateral to the non-implanted ear during binaural stimulation (r = 0.4, p < 0.01; Figure 4A), for both the contralateral and ipsilateral auditory regions during acoustic stimulation (r = 0.63 and r = 0.54, respectively, p < 0.01; Figure 4 B & C), and for the ipsilateral auditory cortex during CI-only stimulation (r=0.43, p < 0.01; Figure 4 D).

**Figure 4.**
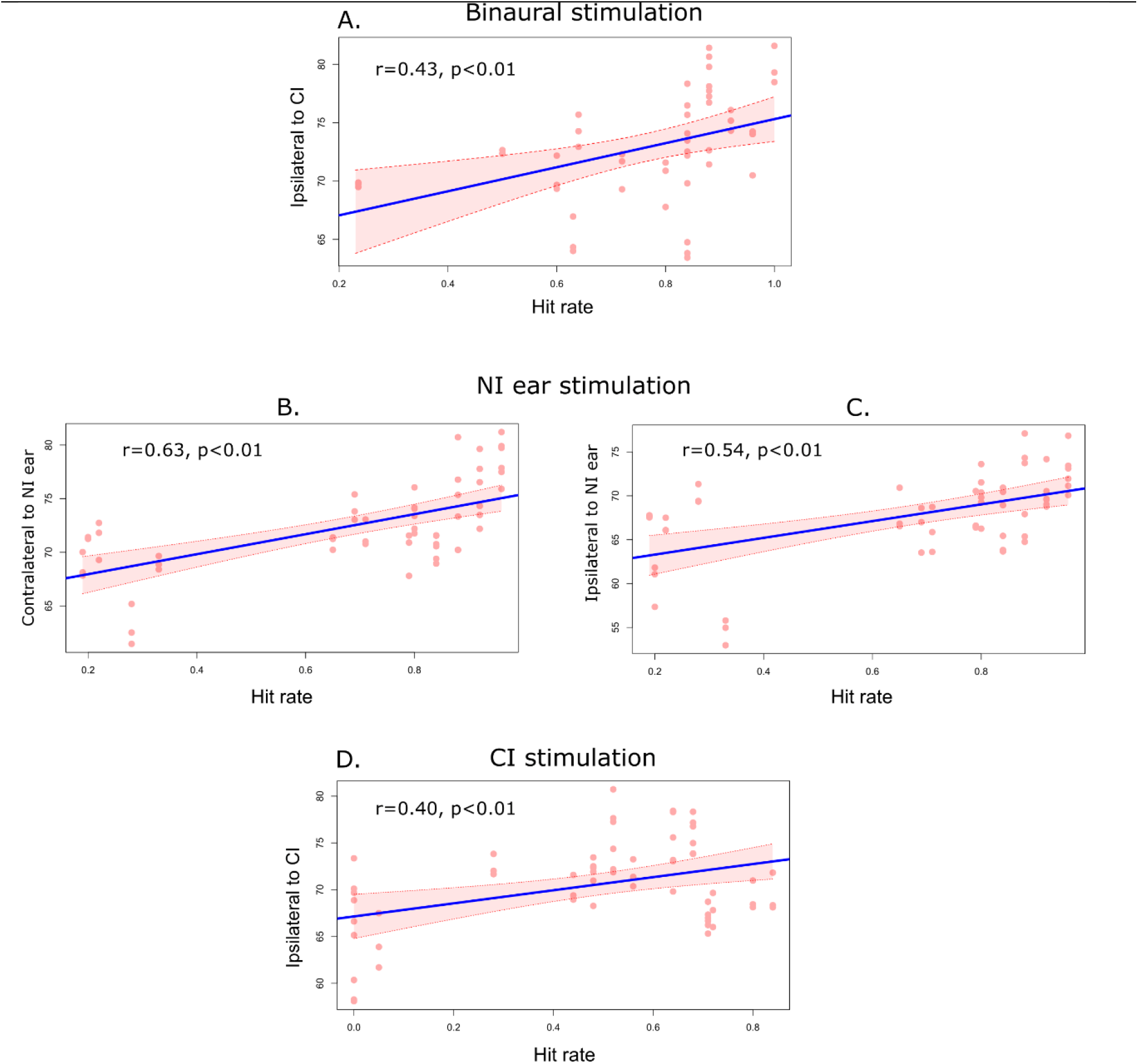
Mean auditory cortical activity and hit rates on the voice/non-voice task. NI ear – non-implanted ear. CI – cochlear implant. There was a significant correlation between the hit rate and activity in the auditory cortex contralateral to the non-implanted ear during binaural stimulation. The highest significant correlations with performance were for the acoustic stimulation (for both auditory cortices). Following CI stimulation, there was a significant correlation between the hit rate and activity in the ipsilateral auditory cortex.

Relationships were also examined between the auditory cortical activity and auditory spatial abilities, expressed by the RMS values on the sound localisation task. A significant correlation was only found for the activity in the hemisphere contralateral to the non-implanted ear (r = 0.3 p < 0.05). This was confirmed by a regression analysis at the whole-brain level between the sound localisation scores and brain activity during binaural stimulation (acoustic and CI stimulation); this pointed to a restricted area located within the contralateral auditory cortex (Figure 5). This result indicates that the auditory cortex (PAC/NPAC) contralateral to the non-implanted ear is implicated in spatial hearing processing in AHL-CI subjects.

**Figure 5.**
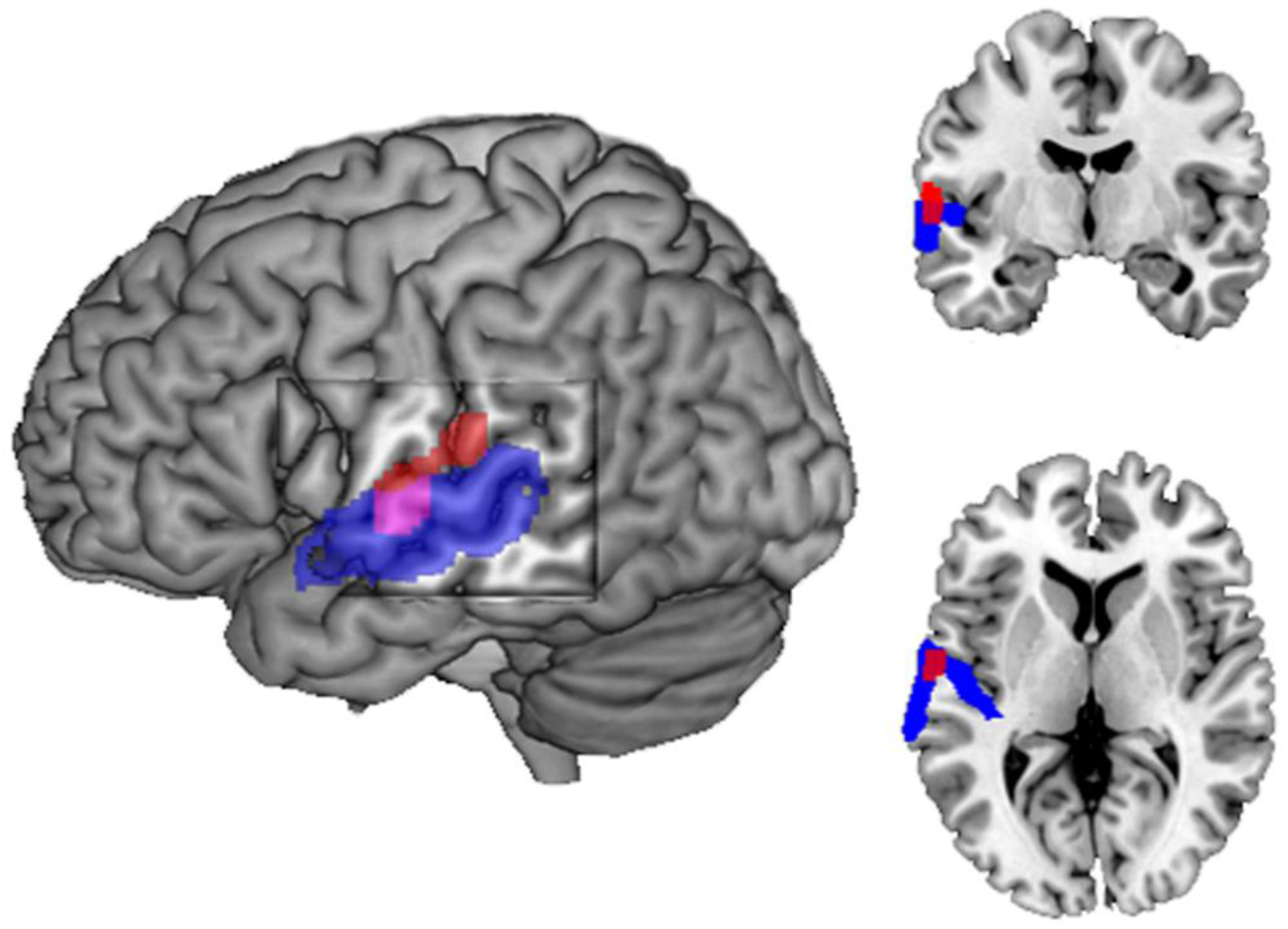
Auditory cortical activity and sound localisation scores (RMS) in patients with a cochlear implant. A whole-brain regression analysis indicates that the auditory cortex contralateral to the non-implanted ear is implicated in spatial processing. The red areas show the location of the significant regression (p corr. <0.05); the blue areas show the cytoarchitectonic auditory cortex.

To summarise, the relationship between the performance levels and the auditory cortical activity was mainly driven by the non-implanted ear during both the acoustic and the binaural conditions. Conversely, the performance levels only related to the CI evoked activity in the auditory cortex ipsilateral to the CI.

#### C. Asymmetries in the auditory activity

The asymmetry index (AI; see materials and methods) indicates which hemisphere is more active according to the side of the auditory stimulation (Figure 6A). AI values are positive when there is greater contralateral activity, and negative when there is greater ipsilateral activity. We found that the control subjects had greater contralateral activity for both of the monaural stimulation conditions; during binaural stimulation, the index did not differ significantly from zero, thus indicating similar levels of activity in both the left and the right auditory cortices (Figure 6A).

**Figure 6.**
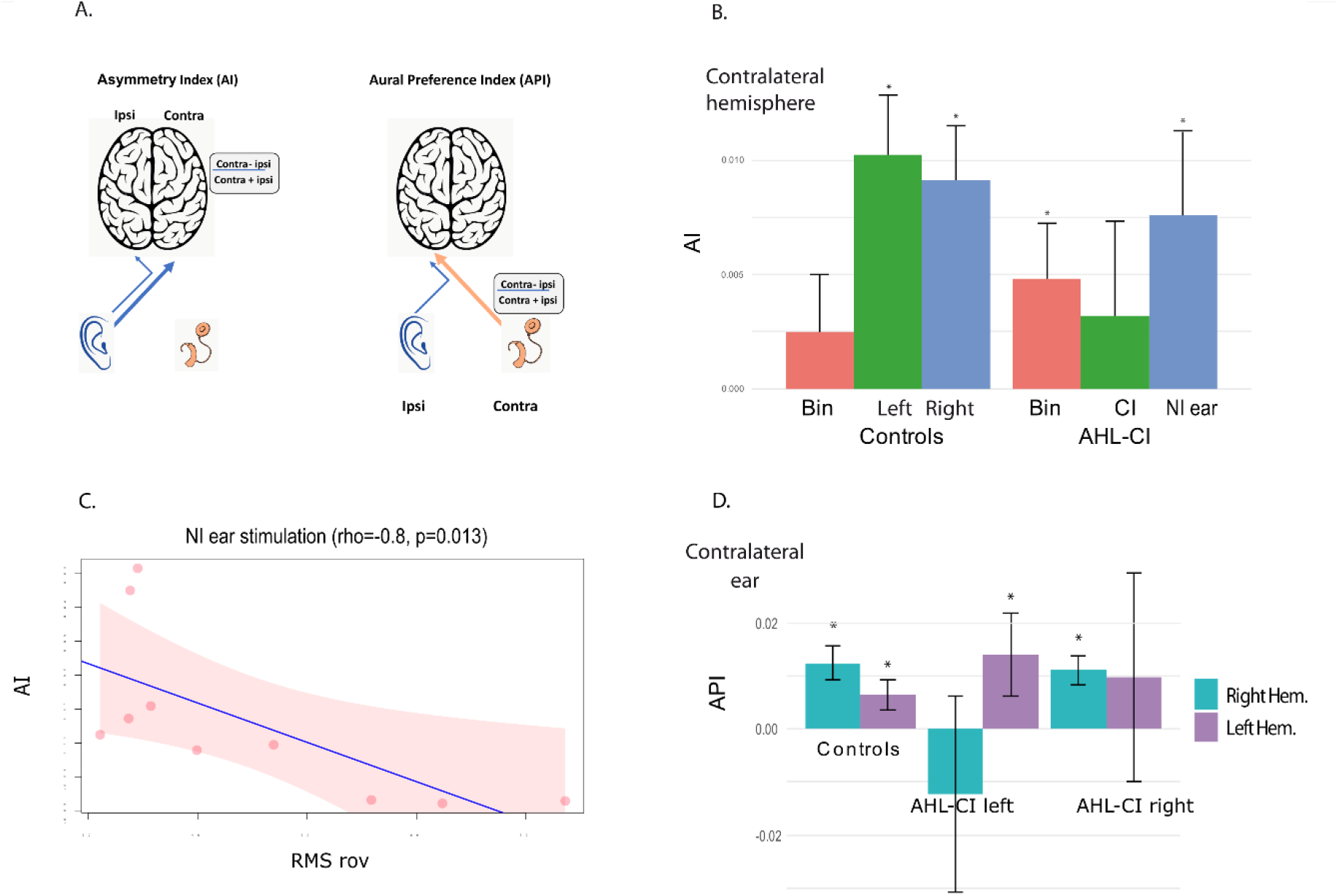
Asymmetry and aural preference indices for the auditory cortex in subjects with a cochlear implant and controls. **A**. Explanatory schemes of the asymmetry index and aural preference index. Asymmetry index (AI) was calculated for each condition using the formula: AI = (Contralateral - Ipsilateral) / (Contralateral + Ipsilateral), where contralateral and ipsilateral refer to the ear stimulated. The aural preference index was calculated using the formula: API = (Contralateral ear – Ipsilateral ear) / (Contralateral ear + Ipsilateral ear). **B**. AHL-CI – patients with a cochlear implant for asymmetric hearing loss. AI – asymmetry index. NI ear – non-implanted ear. Bin – binaural stimulation. CI – cochlear implant stimulation. RMS rov – root mean square scores on the roving condition of the sound localisation task. For binaural stimulation, the asymmetry index in patients was calculated with respect to the non-implanted ear and in controls with respect to the right ear. AI is marked with an asterisk when significantly different from zero (bootstrap, p < 0.05). In AHL-CI patients, there were significant positive (contralateral) AI values for stimulation of the non-implanted ear, as seen in normally-hearing subjects. For stimulation through the CI, the AI index was not significantly different from zero; and for binaural stimulation, the AI index showed a contralateral shift with respect to the non-implanted ear. **C**. The AI for acoustic stimulation correlated highly with the sound localisation scores, implying that smaller errors in spatial localisation were related to the restoration of contralateral hemispheric dominance. **D**. AHL-CI – patients with a cochlear implant for asymmetric hearing loss. API – aural preference index. The API is marked with an asterisk when significantly different from zero (bootstrap, p < 0.05). In normally-hearing controls, a contralateral aural preference was seen for both auditory cortices (API values significantly different from zero). In the left-implanted subjects, there was a contralateral right aural preference for the left auditory cortex; in the right-implanted participants, there was a left aural preference for the right auditory cortex.

In the AHL-CI subjects, there were significant positive AI values when the non-implanted ear was stimulated (Figure 6B). This demonstrates that the acoustic stimulation induced a contralateral dominance, as observed in normally-hearing subjects. Following stimulation through the cochlear implant, the AI index values also tended to be positive, but this was not statistically significant (bootstrap, p > 0.05). The AI differences between the AHL-CI group and the controls were not found to be statistically significant for any of the stimulation conditions.

These results indicate that cochlear implantation restores the normal pattern of interhemispheric asymmetry, with a reactivation of the contralateral auditory cortex in response to stimulation at the non-implanted ear. This can be attributed to the partial recovery of binaural hearing through cochlear implantation. As discussed previously, UHL patients display ipsilateral activation in response to auditory stimulation at the better-hearing ear (Vannson et al., 2020). In contrast, the AHL-CI patients displayed contralateral activation that was similar to the pattern seen in normally-hearing controls.

We also examined whether the AHL-CI subjects’ asymmetry index related to performance on the sound localisation task. We found that the AI following stimulation of the non-implanted ear was highly correlated (rho = -0.8, p = 0.0014) with the localisation performance, i.e., RMS errors in the roving condition. Superior spatial performance therefore related to the restoration of a normal pattern of interhemispheric asymmetry (Figure 6C).

#### D. Aural preference indices of cortical activity

The aural preference index (API; see materials and methods) is used to assess the respective influence of the contralateral and ipsilateral ear on the auditory cortical activity (Kral et al., 2013) (Figure 6A). API values are positive when the contralateral ear has a greater influence. For the control subjects, we found that the API was high for both auditory cortices, with values that differed significantly from zero (p < 0.05; Figure 6D). For the AHL-CI subjects with a left-sided cochlear implant, there was a significant contralateral aural preference for the left auditory cortex (API of about 1%), reflecting a preference for the non-implanted right ear; for the right auditory cortex, the API values did not differ significantly from zero, thus implying that the implanted ear did not strongly influence the contralateral hemisphere (Figure 6D). For the patients with a right-sided implant, the pattern of results was similar, with a significant contralateral aural preference for the right auditory cortex, but not for the left auditory cortex (Figure 6D). These results imply that the hemisphere contralateral to the non-implanted ear shows a contralateral aural preference, similar to that seen in controls.

The API values for the left-implanted and right-implanted AHL-CI patients were pooled into a single group. Correlation analyses showed that the longer the duration of the deafness, the stronger the contralateral aural preference for the auditory cortex ipsilateral to the implant (rho = 0.67, p < 0.05).

## Discussion

This study aimed to explore how the restoration of binaural hearing in AHL through cochlear implantation can restore both spatial hearing processing and a normal pattern of brain activity in auditory areas. We used a sound localisation task and PET brain imaging to study a group of AHL-CI subjects and matched controls. The results provide strong evidence that the restoration of binaural inputs re-establishes the physiological lateralisation of auditory cortical activity and that this relates to the recovery of auditory spatial abilities.

### Partial restoration of spatial hearing after CI

Unilateral or asymmetric hearing loss greatly affects the auditory processes involved in spatial hearing. For instance, AHL impedes the analysis of binaural disparities, which leads to deficits in sound localisation and speech comprehension in noisy environments (review in (Kumpik and King, 2019)). In this study, we showed that cochlear implantation reintroduces binaural indices, leading to improved sound localisation in AHL. We found a substantial binaural benefit (20-30%) for spatial performance (lower RMS), with better scores on the CI-ON condition compared with the CI-OFF condition. However, the performance remained impaired and was particularly low when the stimulus amplitude was roved, with the difference between the CI-OFF and CI-ON conditions becoming only a tendency. Nevertheless, our results suggest that AHL-CI subjects learn to use binaural cues, most probably related to interaural level differences (Seeber et al., 2004; Távora-Vieira et al., 2015) in a head shadow-like effect (Wanrooij and Opstal, 2004), which become ineffective in the amplitude roving condition. These results provide further evidence that AHL-CI subjects can recover substantial spatial hearing abilities when binaural inputs are restored (Dorman et al., 2016; Legris et al., 2020; Litovsky et al., 2019; Polonenko et al., 2018; Zeitler et al., 2015), even though the electrical information is distorted and temporally offset (Zirn et al., 2015).

### Restoration of cortical activity after CI

In adult subjects with acquired AHL, acoustic stimulation of the deaf ear leads to lower levels of cortical activity compared with normally-hearing subjects, depending on the level of the hearing loss (Burton et al., 2012; Langers et al., 2005; Vannson et al., 2020). In the present study, we showed that auditory input through a CI was able to restore cortical activity. This has also been shown in studies on CI-recipients with bilateral profound deafness, where there is evidence for progressive reactivation of the auditory cortex in response to natural sounds as well as reactivation of the neural networks involved in language processing (Giraud et al., 2001; Green et al., 2005; Mortensen et al., 2006; Naito et al., 1995; Stropahl and Debener, 2017). The rather strong activity observed in both hemispheres following CI stimulation in our study (about 80% of the controls) related to the amount of CI experience and the duration of the hearing loss, as is classically reported after CI rehabilitation (Giraud et al., 2001; Rouger et al., 2011; Strelnikov et al., 2015, 2013). However, overall, we found a lower level of auditory cortical activity compared with controls, which may reflect a weaker efficiency of the electric stimulation following a relatively restricted period of activation (Kral et al., 2019, 2006). This may also reflect a lower response to the non-implanted ear, as the AHL-CI subjects had a certain amount of hearing loss in this ear (PTA 15–68 dB); there may also be fewer cortical interactions between the inputs from each ear, as the activity levels for acoustic and binaural stimulation depended on the duration of CI experience.

### Restoration of the functional lateralisation of cortical activity following CI

The contralateral lateralisation of auditory information processing is present at all levels of the auditory pathway (Schönwiesner et al., 2007) and is referred to as the contralateral ear dominance. The disruption of binaural inputs in unilateral or asymmetric hearing loss leads to a shift in hemispheric dominance ipsilateral to the better-hearing ear. This has been found in humans (Burton et al., 2012; Langers et al., 2005; Ponton et al., 2001; Scheffler, 1998; Vannson et al., 2020) and also in animal models of both congenital (Kral et al., 2013; Tillein et al., 2016) and acquired unilateral deafness (McAlpine et al., 1997; Popelár et al., 1994).

It has been suggested that the lateralisation of auditory information processing may relate to the contralateral sound field rather than the contralateral ear *per se* (Eisenman, 1974; Middlebrooks and Pettigrew, 1981; Phillips and Gates, 1982). This hypothesis was originally proposed because of the inhibitory influence of ipsilateral stimulation on auditory cortical neurons (Brugge and Merzenich, 1973; Phillips and Irvine, 1981, 1979) and the low proportion of such neurons unresponsive to ipsilateral stimulation (7% according to (Hall and Goldstein, 1968)). The theory has been supported by several unilateral lesion studies which showed that sound localisation was mainly impaired in the sound field contralateral to the lesion, irrespective of whether the lesion affected a lower or higher stage of brain processing (see (Thompson and Masterton, 1978) for the brainstem; (Sanchez-Longo and Forster, 1958) for the temporal lobe; (Clarey et al., 1992) for a review). However, whether it reflects ear dominance or the sound field, it is clear that the usual pattern of lateralisation is disrupted following unilateral and asymmetric hearing loss. The first aim of our study was to determine whether cochlear implantation could restore this functional lateralisation in AHL.

The aural preference index (API, see Figure 6D) is a commonly used quantitative measure to assess functional lateralisation. It has been used in both human (Gordon et al., 2015) and animal studies (Kral et al., 2013), and has characterised *Aural Preference Syndrome*, which can occur in unilateral deafness (Gordon et al., 2015). API values reflect the strength of activation resulting from each ear at the level of a single hemisphere. In normally-hearing controls, the API reveals a contralateral aural preference for both hemispheres (Figure 6D); in unilateral and AHL, this preference shifts towards the better-hearing ear for both hemispheres (Kral et al., 2013). In our study, we showed that contralateral aural preference is not restored following cochlear implantation in AHL, as CI stimulation activates the contralateral auditory cortex as much as the ipsilateral non-implanted ear (see Figure 6D). These results are in agreement with a study on AHL-CI children (Lee et al., 2020; Polonenko et al., 2018), where there was only a slight, non-significant tendency towards a contralateral preference for the implanted ear.

It is important to be aware that the API compares the level of cortical activation resulting from two auditory pathways (from the cochlea to the cortex) that differ in their functional integrity. It is known that deafness leads to a structural alteration of the anatomical pathway (Eggermont, 2017; Moore et al., 1994; Syka, 2002), and this, coupled with the degraded sound representation provided by the neuroprosthesis (Sato et al., 2017), could weaken the efficiency of the cochlear implant, leading to a weak or absent contralateral aural preference for the implanted ear. However, the contralateral aural preference for the non-implanted ear is preserved, both before and after cochlear implantation (present study and (Lee et al., 2020)). When considering these results, it should be noted that the API does not reflect functional lateralisation concerning representations of the contralateral sound field.

Another quantitative index, the asymmetry index (AI; or lateralisation index in (Vannson et al., 2020) (see Figure 6) is also frequently used to assess auditory cortical processing in normal hearing (Jamison et al., 2006; Schönwiesner et al., 2007; Stefanatos et al., 2008) as well as in deafness and its treatment (Lee et al., 2020). The AI provides a direct comparison of the strength of activation in the auditory cortical areas of the two hemispheres following stimulation at a single ear. The lateralisation in normally-hearing controls is generally reported to be contralateral, meaning that the auditory pathways only weakly activate the auditory cortex ipsilateral to the stimulated ear. This weak ipsilateral activation is partly due to the small number of projections in the ipsilateral pathways (Schönwiesner et al., 2007) and partly due to the various excitatory/inhibitory ipsilateral inputs that reach the auditory cortex (see (Gutschalk and Steinmann, 2015)). These neuronal interactions are important because they lead to contralateral sound field representation (Clarey et al., 1992; Phillips and Gates, 1982; Samson et al., 2000), which is thereby reflected in the AI values.

In a previous study on adult subjects with AHL (Vannson et al., 2020), we demonstrated that the AI shifts towards the ipsilateral auditory cortex following stimulation of the better-hearing ear. This leads to disruption of the neuronal interactions in this auditory cortex. We found that the AI shift was related to the deficit in sound localisation, thus suggesting that a change in activation towards the ipsilateral auditory cortex disrupts representations of the sound field. In the present study, we present data that mirrors this finding, showing that the restoration of auditory input to the deaf ear through the CI re-establishes the contralateral dominance for the better-hearing ear. The was also a trend towards contralateral dominance for the implanted ear, although this was not statistically significant. In addition, the extent of this contralateral lateralisation was found to be associated with better performance on the sound localisation task, thus suggesting a normalisation of the cortical representation of the sound field. This strengthens our previous claim that the asymmetry index can predict the recovery of spatial hearing.

At present, there is no clear evidence concerning the mechanisms that could underlie the loss and the recovery of contralateral activation in AHL and AHL-CI subjects, respectively. It is possible that there are changes in the balance of excitatory and inhibitory interactions between the ipsilateral and contralateral inputs; this may also affect the spatial receptive fields for auditory localisation (Imig et al., 1990).

### Brain plasticity in the primary auditory cortex in deafness and its rehabilitation

Only a few studies have investigated whether the reactivation of a deaf ear can counteract the cortical changes induced by AHL (reviewed by (Vanderauwera et al., 2020). However, none of these studies established a link between brain plasticity and performance on auditory tasks, as we have shown here. In addition, some of the studies only investigated single cases (Firszt et al., 2013; Sharma and Glick, 2016), while others had highly variable data (Lee et al., 2020), thus precluding any robust conclusions. Most of these previous studies used EEG to study the neural changes (Lee et al., 2020; Legris et al., 2020, 2018; Polonenko et al., 2018), which limits the spatial resolution of any findings. They were therefore unable precisely locate the functional reorganisation that takes place, as EEG relies on algorithms of source reconstruction, which lack accuracy. In our study, the PET methodology was able to precisely locate the restoration of functional lateralisation to the auditory cortex (primary and non-primary). It was also possible to show that the auditory cortical activity relates to sound localisation performance. It was also possible to show, using a whole-brain regression analysis, that the function integrity (capacity to respond) of the auditory cortex is predictive of auditory spatial perception. These data highlight the early stages of auditory processing in the cortex as a key level that links brain plasticity to spatial hearing skills. However, brain plasticity may also take place in areas outside of the auditory cortex, such as when acoustical and electrical information are combined to process non-spatial auditory information (Strelnikov et al in preparation).

## Conclusion

In this study, we demonstrated that the restoration of binaural stimulation in AHL-CI patients reverses the abnormal lateralisation pattern induced by AHL. In addition, the hemispheric dominance contralateral to the non-implanted ear was found to be associated with superior sound localisation. This supports the hypothesis that aural dominance relates to representations of the spatial sound field; this would be disrupted in unilateral deafness and restored in AHL-CI patients. The results suggest that cochlear implantation rehabilitates the binaural excitatory/inhibitory cortical interactions, and that these enable the recovery of the spatial selectivity involved in sound localisation.

## Data Availability

All data produced in the present study are available upon reasonable request to the authors

## Acknowledgment

We thank all of our subjects for their participation in the study, and the ENT team at Purpan hospital, Toulouse, for their precious help and support. The authors would also like to acknowledge all the technical team of the PET scan platform for their help during the scanning sessions, especially Mathieu Alonso, Kevin Rouchette for the technical development of the PET study, and Dr. Anne Hitzel, Dr. Anne Julian for medical support of the PET study. The authors are very thankful to Jessica Foxton for correction and critical review of the article.

